# Pre-exposure prophylaxis with Evusheld™ elicits limited neutralizing activity against the omicron variant in kidney transplant patients

**DOI:** 10.1101/2022.03.21.22272669

**Authors:** Ilies Benotmane, Aurélie Velay, Olivier Thaunat, Gabriela Gautier Vargas, Jérôme Olagne, Samira Fafi-Kremer, Sophie Caillard

## Abstract

The combination of cilgavimab-tixagevimab (Evusheld™, Astra Zeneca) became the mainstay for protecting transplant recipients with poor response to vaccination against the omicron variant. Serum neutralizing capacity against SARS-CoV-2 is positively associated with protection against severe forms of Covid-19.

Both anti-RBD IgG titers and neutralizing antibody titers against the omicron BA.1 variant were measured in serum samples collected from 63 adult kidney transplant recipients who received prophylactic injections of Evusheld™. Patients who received prophylactic Ronapreve™ (casirivimab-imdevimab, n = 39) and those who were infected with SARS-CoV-2 during the fifth wave of the pandemic (n = 14) served as negative and positive controls, respectively.

After a median interval from injection of 29 days (interquartile range 29-33 days), only 9.5% of patients who received Evusheld™ were able to neutralize the omicron variant compared to 71% of patients who were infected with SARS-CoV-2 and 2.6% of those who received Ronapreve™. Interestingly, convalescent patients displayed higher levels of neutralizing antibodies than those who received EvusheldTM (median: 2.3 log IC50, IQR: 1.5-2.7 versus 0.00 log IC50, IQR: 0&[ndash] 0.05; p<0.001). A high interindividual variability in anti-RBD IgG titers was observed after Evusheld™ (range: 262-7032 BAU/mL). This variability was largely explained by the patients’body mass index, which showed an inverse correlation with anti-RBD IgG titers.

These findings suggest that Evusheld™ given at a dose of 300 mg is not sufficient to elicit an anti-RDB titer that confers in vivo neutralizing activity and support recent FDA recommendations, derived from in vitro models, regarding the need to increase the dose of Evusheld™

## Introduction

Kidney transplant recipients show an impairment of vaccine-induced immune response, resulting in an increased risk of severe Covid-19 (1). In an effort to address this issue, health authorities have authorized the use of anti-SARS-CoV-2 monoclonal antibodies for pre-exposure prophylaxis. While the combination of casirivimab–imdevimab (Ronapreve™, Roche Regeneron) has been shown to confer satisfactory protection against the delta variant (2), it has limited neutralizing activity against omicron (3). Consequently, as of December 2021, the combination of cilgavimab–tixagevimab (Evusheld™, Astra Zeneca) became the mainstay for protecting transplant recipients with poor response to vaccination against the omicron variant. Serum neutralizing capacity against SARS-CoV-2 is positively associated with protection against severe forms of Covid-19, including the kidney transplant recipients population (4).

### Patients and Methods

Both anti-RBD IgG titers and neutralizing antibody titers against the omicron BA.1 variant were measured in serum samples collected from 63 adult kidney transplant recipients who received intramuscular prophylactic injections of Evusheld™ (150 mg tixagevimab and 150 mg cilgavimab) in the Lyon and Strasbourg University Hospitals. Cases with a history of Covid-19 or positive anti-nucleocapsid IgG were excluded. Patients who received prophylactic Ronapreve™ (600 mg casirivimab and 600 mg imdevimab, n = 39) and those who were infected with SARS-CoV-2 during the fifth wave of the pandemic (n = 14) served as negative and positive controls, respectively. Ethical approval for this study was obtained from the local Ethics Committees (identifiers: DC-2013-1990, Lyon University Hospital and CE-2021-9, Strasbourg University).

## Results

After a median interval from injection of 29 days (interquartile range [IQR]: 29-33 days), patients who received Evusheld™ had a low neutralizing activity (Figure 1A) and only 9.5% of them (6/63) were able to neutralize the omicron variant compared to 71% of patients [10/14] who were infected with SARS-CoV-2 and 2.6% [1/39] of those who received Ronapreve™. Interestingly, convalescent patients displayed higher levels of neutralizing antibodies than those who received Evusheld™ (median: 2.3 log IC50, IQR: 1.5-2.7 *versus* log IC50, IQR: 0-0.05; p<0.001). While anti-RBD IgG titers were generally low after Evusheld™ injection (median: 2583 binding antibody unit (BAU)/mL, IQR: 1906-3611 BAU/mL), a high interindividual variability was observed (range: 262-7032 BAU/mL, Figure 1B). This variability was largely explained by the patients’ body mass index, which showed an inverse correlation with anti-RBD IgG titers (Figure 1C). Further analysis revealed that patients with anti-RBD titers <2500 BAU/mL after Evusheld™ injection had no neutralizing activity (Figure 2). Furthermore, seven patients in this cohort developed symptomatic Covid-19 including two who required hospitalization. All had negative neutralizing activity at the time of diagnosis.

**Figure 1A.**
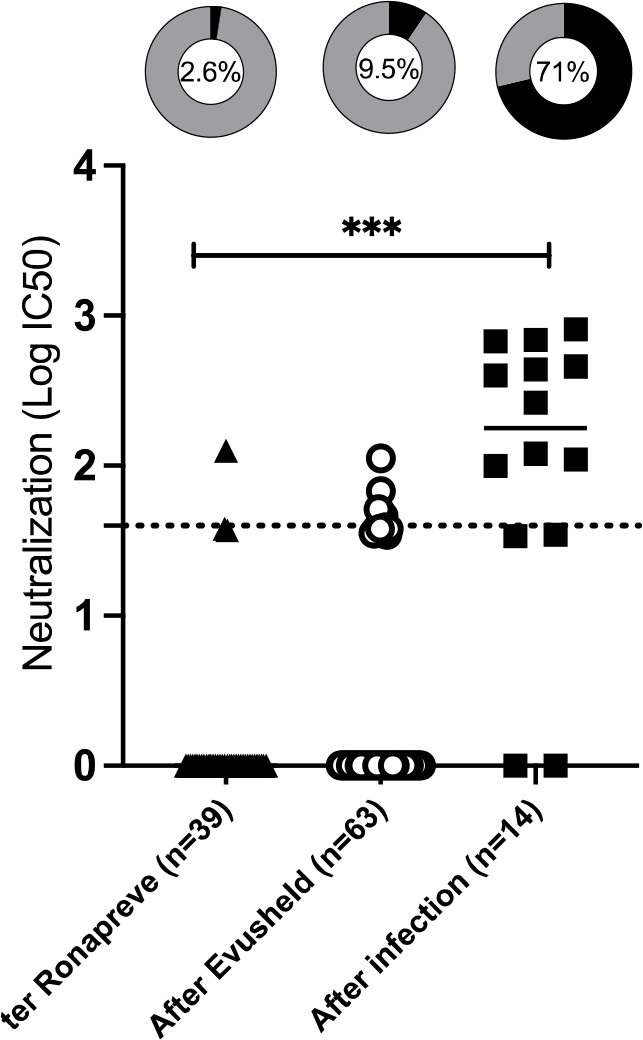
Serum neutralizing IgG titers (log IC50) measured with an in-house viral pseudoparticle-based assay described before (4) in three groups of kidney transplant recipients. Circles denote titers measured at 28 post-injection days in patients (n = 63) who received Evusheld™ (300 mg), whereas triangles indicate titers quantified at 31 post-injection days in patients (n = 39) who received Ronapreve™ (1200 mg). Squares denote titers measured at 27 post-infection days in patients (n = 14) who were infected with SARS-CoV-2. Dotted line represents the neutralizing positivity threshold (1.6 log IC50). Groups were compared with the Kruskal-Wallis test. The contingency graphs at the bottom of the figure indicate the percentages of patients with neutralizing activity in each group with (positive in black and negative in gray; the percentage is reported in the middle).

**Figure 1B.**
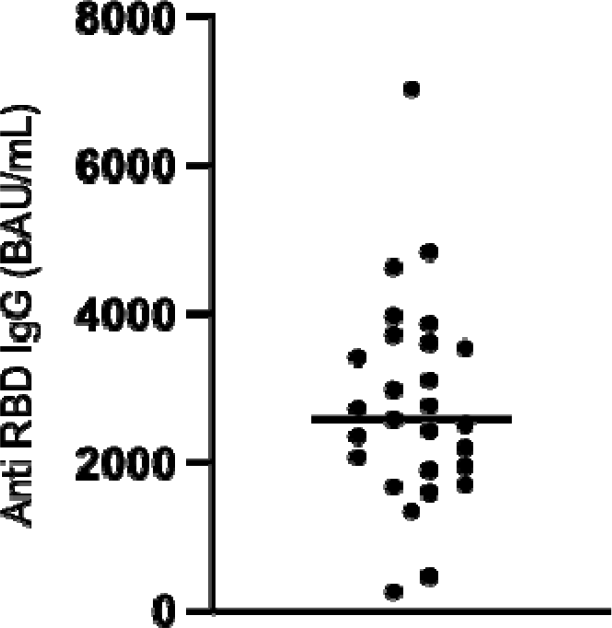
Anti-RBD IgG titers (BAU/mL, Abbott Architect, Chicago IL, USA) 28 days after Evusheld™ injection (300 mg) in 27 patients who did not receive Ronapreve™ before Evusheld™.

**Figure 1C.**
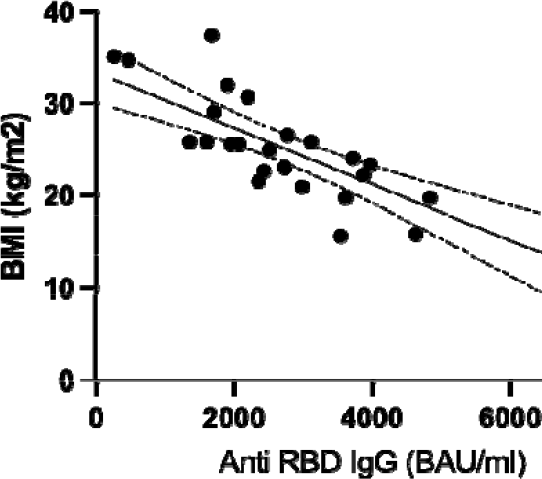
Correlation between body mass index (kg/m^2^) and anti-RBD IgG titers (BAU/mL, Abbott Architect, Chicago IL, USA) measured at 28 days after Evusheld™ injection (300 mg) in 27 patients who did not receive Ronapreve™ before Evusheld™; r^2^=0.595.

**Figure 2.**
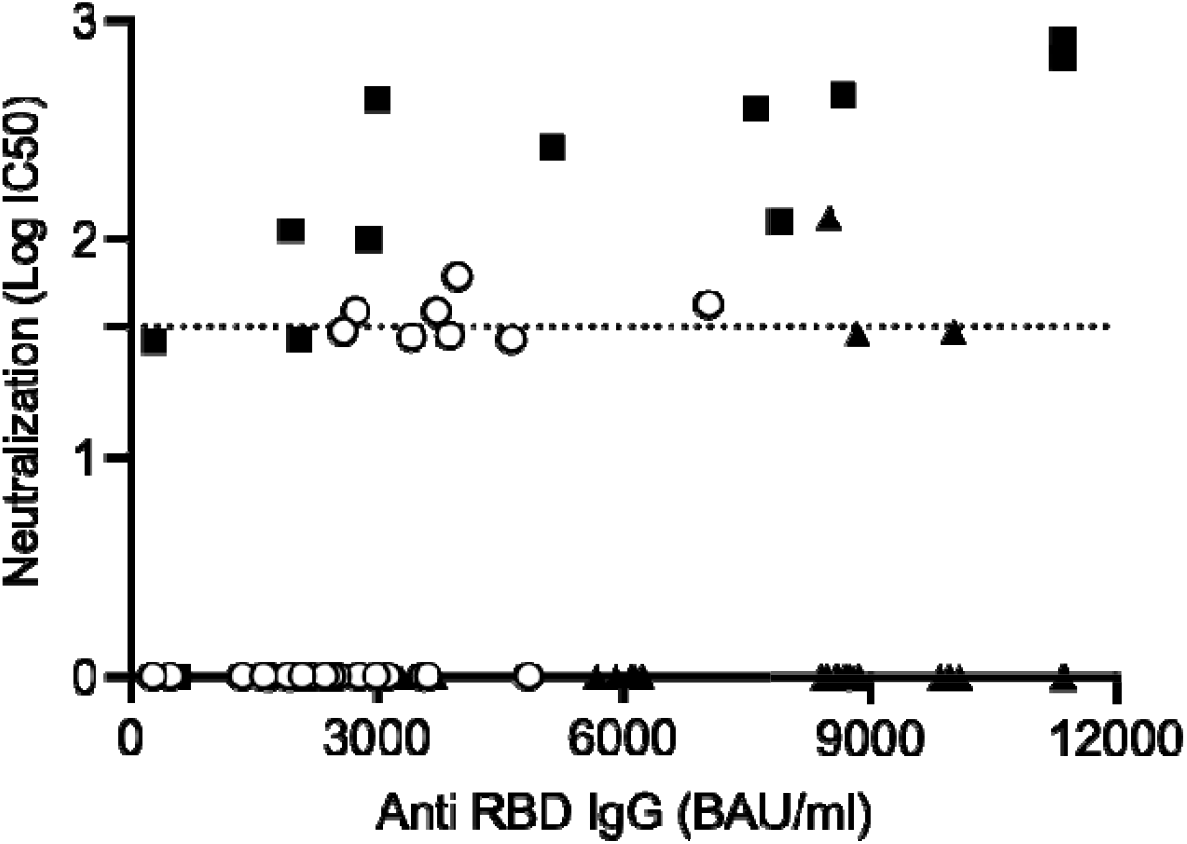
Correlation between anti-RBD IgG (Abbott Architect, Chicago IL, USA) and neutralizing antibody titers (3) in three groups of kidney transplant recipients. Circles denote titers measured at 28 post-injection days in patients (n = 63) who received Evusheld™ (300 mg), whereas triangles indicate titers quantified at 31 post-injection days in patients (n = 39) who received Ronapreve™ (300 mg). Squares denote titers measured at 27 post-infection days in patients (n = 14) who were infected with SARS-CoV-2.

## Discussion

The results of our study indicate that less than 10% of patients who received Evusheld™ were able to neutralize the omicron BA.1 variant at 29 post-injection days. These findings suggest that Evusheld™ given at a dose of 300 mg is not sufficient to elicit an anti-RDB titer that confers *in vivo* neutralizing activity. This is in line with a recent study showing that anti-RBD levels of approximately 8500 BAU/mL were associated with serum neutralizing activity against omicron in transplant recipients who received three vaccine doses (5). Our data also support recent FDA recommendations (6), derived from *in vitro* models, regarding the need to increase the dose of Evusheld™.

## Data Availability

All data produced in the present study are available upon reasonable request to the authors

## References

1. Caillard S, Thaunat O. COVID-19 vaccination in kidney transplant recipients. Nat Rev Nephrol. 2021 Dec;17(12):785–787.

2. Ducloux D, Courivaud C. REGEN-Cov antibody combination to prevent COVID-19 infection in kidney transplant recipient without detectable antibody response to optimal vaccine scheme. Kidney Int. 2022;101(3):645–646.

3. Planas D, Saunders N, Maes P, et al. Considerable escape of SARS-CoV-2 Omicron to antibody neutralization. Nature. 2022;602(7898):671–675.

4. Charmetant X, Espi M, Benotmane I, et al. Infection or a third dose of mRNA vaccine elicit neutralizing antibody responses against SARS-CoV-2 in kidney transplant recipients. Sci Transl Med. 2022 Feb 1:eabl6141. DOI: 10.1126/scitranslmed.abl6141. Epub ahead of print. PMID: 35103481.

5. Kumar D, Hu Q, Samson R, Ferreira VH, et al. Neutralization against Omicron variant in transplant recipients after three-doses of mRNA vaccine. Am J Transplant. 2022 Mar 10. DOI: 10.1111/ajt.17020. Epub ahead of print. PMID: 35266606.

6. https://www.fda.gov/drugs/drug-safety-and-availability/fda-authorizes-revisions-evusheld-dosing. Last access 24 Feb 2022

